# Effect of rearfoot strikes on the hip and knee rotational kinetic chain during the early phase of cuttings in female athletes

**DOI:** 10.1101/2021.02.22.21251902

**Authors:** Issei Ogasawara, Yohei Shimokochi, Shoji Konda, Tatsuo Mae, Ken Nakata

**Affiliations:** Department of Health and Sports Sciences, Graduate School of Medicine, Osaka University, 1-17 Machikaneyama-cho, Toyonaka, Osaka 560-0043, Japan; Department of Health and Sport Management, Osaka University of Health and Sport Sciences, 1-1 Asashirodai, Kumatori-cho, Sennan-gun, Osaka 590-0496, Japan; Department of Sports Medical Biomechanics, Graduate School of Medicine, Osaka University, 2-2 Yamada-oka, Suita, Osaka 565-0871, Japan

**Author notes:** **Corresponding author:** Issei Ogasawara, Department of Health and Sports Sciences, Graduate School of Medicine, Osaka University, 1-17 Machikaneyama-cho, Toyonaka, Osaka 560-0043, Japan. **Financial disclosure related to research:** None.

**Keywords:** foot strike pattern, rotational destabilization, horizontal-plane kinetic chain, deceleration motion, anterior cruciate ligament injury

## Abstract

**Purpose:** Biomechanical factors affecting horizontal-plane hip and knee kinetic-chain and anterior cruciate ligament (ACL) injury risks during cutting maneuvers remains unclear. This study aimed to examine whether different foot strike patterns alter horizontal-plane hip and knee kinetics and kinematics during a cutting maneuver in female athletes and clarify the individual force contribution for producing high-risk hip and knee loadings.

**Methods:** Twenty-five healthy female athletes performed a 60° cutting task with forefoot and rearfoot first strike conditions. Horizontal-plane hip and knee moment components, angles, and angular velocities were calculated using synchronized data of the marker positions on the body landmarks and ground reaction forces (GRF) during the task. The one-dimensional statistical parametric mapping paired t-test was used to identify the significant difference in kinetic and kinematic time-series data between foot strike conditions.

**Results:** In the rearfoot strike condition, large hip and knee internal rotation loadings were produced during the first 5% of stance due to the application of GRFs, causing a significantly larger hip internal rotation excursion than that of the forefoot strike condition. Dissimilarly, neither initial hip internal rotation displacement nor knee internal rotation GRF loadings were observed in the forefoot strike condition.

**Conclusion:** Rearfoot strike during cutting appears to increase noncontact ACL injury risks as the GRF tend to produce combined hip and knee internal rotation moments and the high-risk lower limb configuration. Conversely, forefoot strike during cutting appears to be an ACL-protecting strategy as the ACL-harmful joint loadings and lower-extremity configurations do not tend to be produced. Thus, improving foot strike patterns during cutting should be incorporated into ACL injury prevention programs.

## Introduction

Knee injuries are the second most occurrence injuries to ankle injuries in sports (Silvers-Granelli et al., 2015). Anterior cruciate ligament (ACL) injuries consist of 20.3% of all these knee injuries (Majewski et al., 2006). Female athletes have 2–4 times higher ACL injury incidence rates than male counter parts (Agel et al., 2005; Arendt and Dick, 1995; Montalvo et al., 2019; Waldén et al., 2010) with females of younger than 25 years old are most likely to sustain an ACL tear (Schilaty et al., 2017). ACL injured people at their younger age results in an increased risk of the early onset posttraumatic knee osteoarthritis and considerable spoil the quality of life (Lohmander et al., 2004). More than 70% of those occur without any direct blow to the knee from others (Boden et al., 2000) during the sharp deceleration motion, such as a cutting or landing in a short period of time from the initial foot contact (Krosshaug et al., 2007). Thus, understanding the mechanisms of noncontact ACL injuries as well as risky and safe movement skills for sharp decelerating motions are essential, especially for young female athletes, to prevent sports-related ACL injuries.

Hip internal rotation combined with knee valgus and internal rotation observed at the time of noncontact ACL injuries was considered as a risky lower limb configuration for ACL injuries (Boden et al., 2000; Ireland, 1999; Quatman and Hewett, 2009). A video analysis study (Koga et al., 2018) reported that hip internal rotation followed by subsequent knee internal rotation was commonly observed in 10 injury cases. A lab-controlled study supported the effect of the horizontal-plane hip configuration on the knee loadings that the hip internal rotation at foot impact is a significant predictor of knee valgus loading during cutting maneuvers in female athletes (McLean et al., 2005; Pollard et al., 2007; Sigward et al., 2015). Collectively, these previous studies suggested that the loss of rotational hip control at initial contact (IC) may indirectly alter the mechanical status of the knee, resulting in high loading on the passive knee structures including the ACL. Therefore, identifying the mechanical and technical factor that induces the horizontal hip and knee rotational instability is very interesting from the perspective of ACL injury risk.

Given the noncontact mechanism of ACL injuries, the ground reaction force (GRF) exerted at the center of pressure (CoP) of the landing foot would be a primary external force which is capable of developing the abnormal hip and knee rotational configurations in this time frame. Previous lab-controlled studies have tried to identify the specific joint loading patterns associated with the risky hip and knee configurations (Havens and Sigward, 2015a; McLean et al., 2005; Pollard et al., 2007, 2018). In the most of those studies, only the resultant moment acting on the joint of interest was evaluated as a measure of joint loading, however, how each of the external force component, such as translational and rotational inertia, gravity, free moment and GRF, developed that resultant moment was not well focused in the context of ACL injury. Specifically, knowing the GRF’s contribution on the final joint kinematics may provide an insight into the mechanical cause of the hip and knee kinetic chain observed in the situation of the noncontact ACL injury.

The rearfoot strike (RFS), as a potential technical factor, has been reported to be frequently associated with noncontact ACL injuries (Boden et al., 2009; Koga et al., 2018; Montgomery et al., 2016; Waldén et al., 2015). Lab-controlled studies have also suggested that the different foot strike pattern (Cortes et al., 2012; David et al., 2017) or foot orientation relative to the floor (Kristianslund et al., 2014; Shimokochi et al., 2016) altered the knee loading pattern during cutting or landing maneuvers. Further, our group recently reported that the GRF acting at the rearfoot is more likely to apply to the combined knee valgus and tibial internal rotation moment in the early phase of cutting maneuvers (Ogasawara et al., 2020). Although the previous studies revealed the link between the foot strike pattern and knee loading pattern, it is still unknown whether different foot strike patterns affect the hip and knee horizontal-plane kinetics and kinematics during the deceleration phase of cutting maneuvers in female athletes.

Therefore, the purpose of this study was to i) clarify the individual external force contributions for producing high-risk hip and knee loadings, and ii) investigate whether the different foot strike patterns, forefoot strike (FFS) and RFS, alter the horizontal-plane hip and knee kinetics and kinematics during a cutting maneuver in female athletes. We hypothesized that i) the moment derived from the GRF, among the other types of external forces, significantly determine the magnitude of the hip and knee resultant moments, and ii) on the basis of the potential risk of RFSs, we hypothesized that the RFS would produce a significantly greater GRF-driven internal rotation moment of hip and knee than FFS, and further this GRF loading would produce a greater hip and knee internal rotation angular excursion and angular velocities than the FFS during the early phase of the cutting maneuver.

## Methods

### Ethics statement

The ethics board approved this experiment (Mukogawa Women’s University, No. 12-13), and written informed consent was obtained from all participants.

### Participants

Twenty-five healthy female collegiate handball players participated in this study. All athletes trained 2−3 hours per day and were familiar with the change of direction movements. Athletes who had a history of severe knee injuries, such as an ACL tear, or minor lower limb trauma, such as an ankle sprain, in the 6 months preceding the date of initiation of the experiment were excluded.

### Procedure

Seventeen reflective markers (diameter, 14 mm) were attached to 17 landmarks (Figure 1A, Table 1) by the same researcher (IO). The participants were instructed to change the running direction with a single step (cutting task) at an angle of 60° (Besier et al., 2001; Ogasawara et al., 2020). The reference cone was placed on the floor at an angle of 60° from the approach line to guide the participant. The test leg was determined by asking the participants which leg was preferred to control the foot-floor contact point at the instance of impact.

**Table 1.** Locations of the reflective body markers on the cutting limb and pelvis.

|  |  |
| --- | --- |
| Toe | On the most anterior point of the toe, over the shoe |
| Medial toe | Medial aspect of the most prominent point of the first MP joint, over the shoe |
| Lateral toe | Lateral aspect of the most prominent point of the fifth MP joint, over the shoe |
| Dorsal foot | Dorsal aspect of the midpoint of the second and third MP joints, over the shoe |
| Medial ankle | On the most prominent point of the medial malleolus |
| Lateral ankle | On the most prominent point of the lateral malleolus |
| Heel | On the most posterior point of the shoe heel and 2 cm above the floor level when the subject is standing stationary |
| Medial knee | On the most prominent point of the medial femoral epicondyle |
| Lateral knee | On the most prominent point of the lateral femoral epicondyle |
| Tibial tuberosity | On the most prominent point and anterior aspect of the tibial tuberosity |
| Great trochanter | On the most prominent point of the great trochanters (both sides) |
| Mid thigh | On the halfway (approximately) point of the anterior aspect of the thigh |
| ASIS | On the most prominent point of the ASISs (both sides) |
| PSISs | On the most prominent point of the PSISs (both sides) |

ASIS, anterior superior iliac spine; PSIS, posterior superior iliac spine; MP, metatarsophalangeal

**Figure 1.**
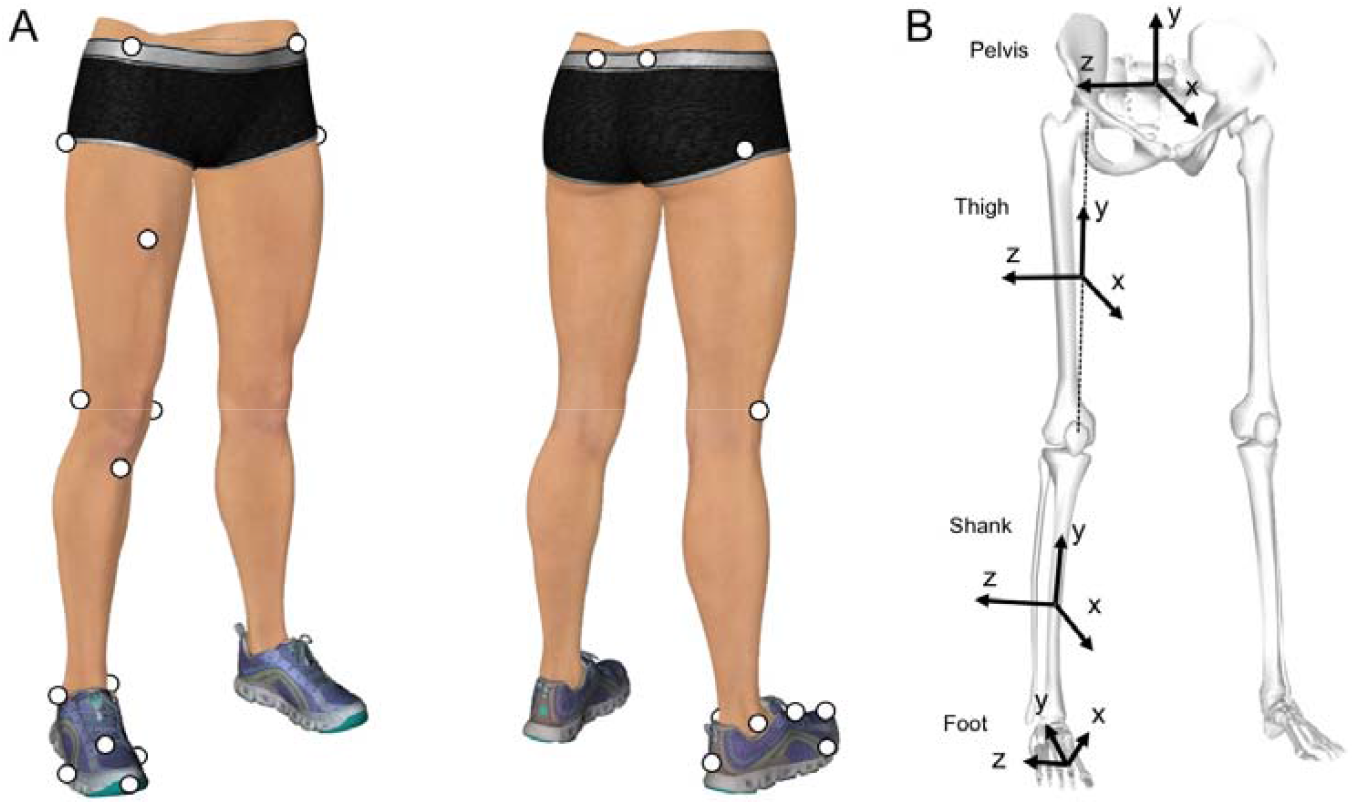
Reflective marker attachment site and orientations of the local coordinate systems. (A) Location of reflective markers on the right leg. (B) The kinematic model consists of 4 segments (foot, shank, thigh, and pelvis) and 3 joints (ankle, knee, and hip). The local coordinate system for each segment is defined as a 3×3 rotation matrix consisting of 3 common perpendicular unit base vectors: the x-axis is the anterior/posterior axis pointing forward, y-axis is the vertical axis along with the longitudinal line of the segment pointing upward, and z-axis is the medial/lateral axis pointing to the right of the segment.

Two different foot strike conditions (RFS versus [vs.] FFS) were tested. In the RFS condition, participants were required to hit the force plate with their heel first to locate the CoP of the rearfoot at the beginning of the stance phase and then move their weight to the forefoot to push off. For the FFS condition, participants were asked to touch the force plate with their forefoot throughout the stance phase. Since we acknowledged that the GRF more frequently applies the combined knee valgus and tibial internal rotation loads when CoP is at the rearfoot (Ogasawara et al., 2020), the approach speed was set at less than 2.0 m/s to reduce the magnitude of the GRF for participants’ safety. The approach speed was monitored online with the data streaming feature of the motion capture system (NatNet, Motive Body 1.1, NaturalPoint, Inc., Corvallis, OR, USA).

After task familiarization was complete, a static standing pose was recorded as the reference for neutral joint angles, and 10 successful trials for each foot condition were recorded. A 3-minute rest between conditions and an approximately 30-second interval between trials were provided to minimize fatigue. To equalize the possible effect of fatigue between foot strike conditions, the order of the 2 conditions was randomized for each participant. The success of the foot strike pattern was visually judged by 2 researchers (IO and assistant YK, CA or KM) for each trial. If a consensus was not achieved, that trial was discarded, and the participant was requested to execute an additional trial. The three-dimensional marker positions were captured using 12 optical cameras (OptiTrack S250e with 250-Hz sampling; software: Motive Body 1.1, NaturalPoint, Inc.) simultaneously with the GRF recordings (force plate: type 9281B, Kistler, Winterthur, Switzerland; data acquisition device: USB-6218 BNC with 1-kHz sampling, National Instruments, Austin, TX, USA). A clock device (eSync, NaturalPoint, Inc.) was used to generate the timing signal to synchronize the onset of the camera and force plate recordings.

### Data processing

The position data were smoothed using a second-order low-pass zero-lag digital Butterworth filter at cut-off frequencies of 12–15 Hz, which were determined by residual analysis (Winter, 2005). The GRF data were smoothed at the same cut-off frequencies as those used for smoothing the marker data (Bisseling and Hof, 2006). The weight acceptance phases (vertical GRF >10 N) of the position and GRF data were extracted and time-normalized (0−100%) throughout the stance phase. The kinematic model was consisted of 4 segments (foot, shank, thigh, and pelvis) with 3 joints (ankle, knee, and hip) (Figure 2). The ankle and knee joint centers were calculated as the midpoint between medial and lateral markers for the ankle and knee joints. The hip joint center was calculated using a previously reported method (Bell et al., 1990). The local coordinate system (LCS) of each segment was defined as a 3 × 3 rotation matrix consisting of common perpendicular unit base vectors where the x-axis (*e*_*i,x*_) was the anterior/posterior axis pointing to the anterior of the segment, y-axis (*e*_*i,y*_) was the vertical axis along with the longitudinal line of the segment pointing upward, and z-axis (*e*_*i,z*_) was the medial/lateral axis pointing right of the body (Grood and Suntay, 1983; Wu et al., 2002). The segmental mass, position of mass center, and inertia moment were estimated based on the data of Japanese athletes (Ae et al., 1992). The hip and knee internal(+) / external(−) rotations were defined as the segmental rotation around the y-axis of the thigh and shank segments, respectively, and those angles were calculated using Derrick et al.’s (Derrick et al., 2020) method for the hip joint and Grood and Suntay’s method (Grood and Suntay, 1983) for the knee joint. The calculated internal/external rotation angles were offset with the angle at the static pose. In addition, the internal/external rotation angular excursions were calculated by subtracting with the angle at IC of each trial to quantify the angular displacements that occurred after foot contact. The internal/external rotational joint angular velocities of the hip and knee were obtained by using the numerical differentiation of the joint angle. See Supplemental Content for the detail of joint angle and angular velocity calculation.

**Figure 2.**
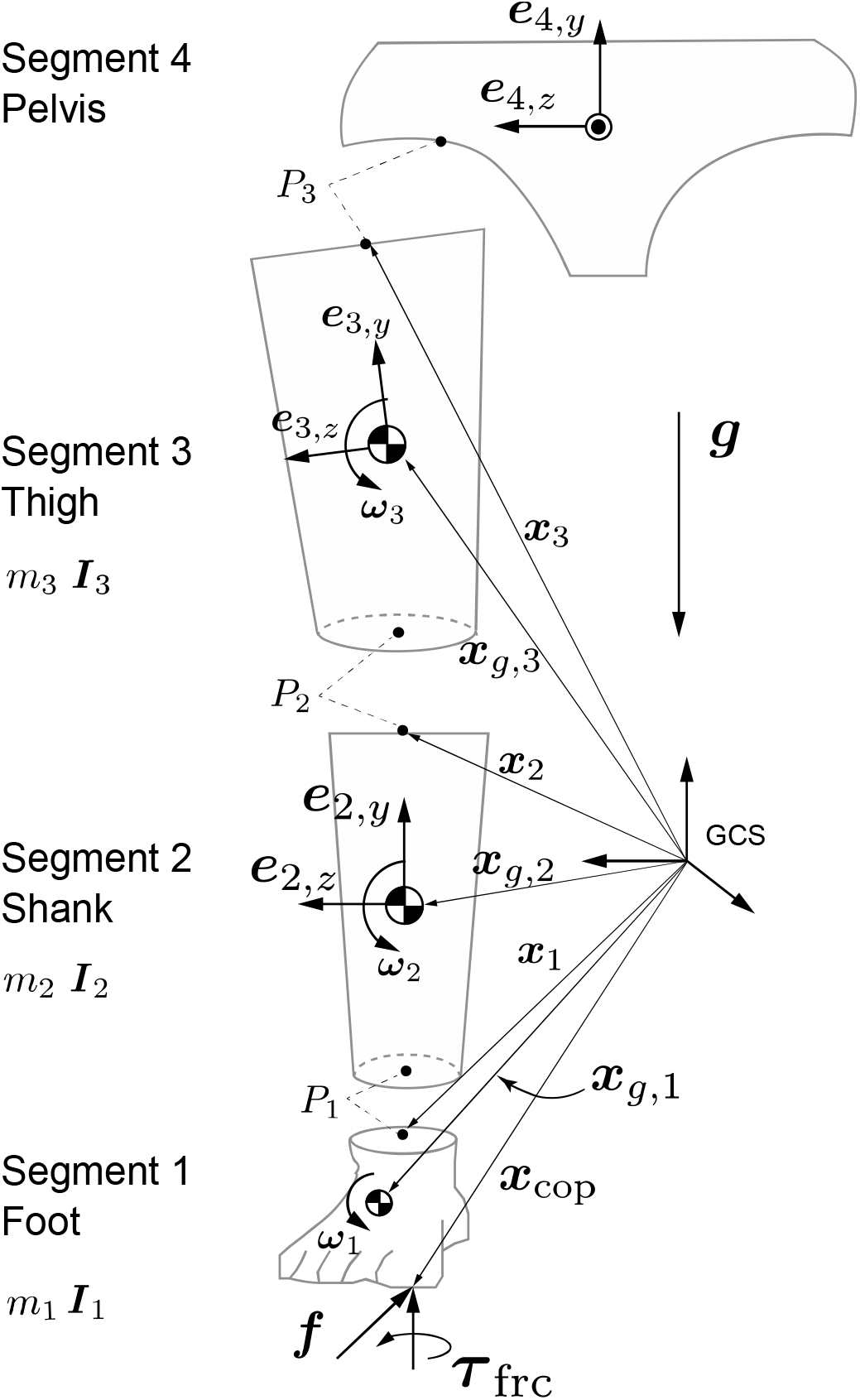
Three-dimensional four-link kinematic model. Schematic frontal view of the kinematic model used in this study. For visibility, each segment is visualized separately at the joint, but they are actually connected.

The Newton-Euler equation of motion was solved to obtain all of the moment components, e.g., 1) resultant moment, 2) moment of GRF, 3) rotational inertia moment, 4) gyroscopic moment, 5) moment of linear inertial force, 6) moment of gravity acting at the segmental mass center, and 7) joint moment due to friction moment acting around the vertical axis at CoP (See Figure s1 in Supplemental Content for the free-body-diagram and the mathematical detail of the inverse dynamics model).

For each moment component, the projections onto the y-axis of the thigh and shank segments were calculated to extract the internal(+) / external(−) rotation components. This process corresponded to express the resultant moment in the LCS of the distal segment (Derrick et al., 2020). According to the joint structure, the resultant hip internal/external rotation moment was assumed to be produced mainly by the hip rotators to balance the external loadings. In contrast, the resultant knee internal/external rotation moment was regarded as a resistive moment derived from the stretched passive structures including the ACL (Derrick et al., 2020), under the presence of the external loadings on the knee. All of the numerical calculations were done with the custom scripts of Scilab 6.0.0 (https://www.scilab.org/).

### Statistical analysis

An a priori power analysis using G*Power 3.1.9.4 suggested that the appropriate sample size for the paired t-test was n=22 to achieve 80% power at a statistical significance criterion of 0.01, with a large effect size (ES) (Cohen d >0.8).

To confirm whether the approach speeds differed between foot strike conditions, the average speeds of the midpoint between both anterior superior iliac spine markers from −40 to 0 ms before foot contact were compared using the paired t-test (P<0.05).

To examine the contribution of each moment component on the magnitude of the resultant hip and knee moments, the time-series data of each moment component was normalized by the resultant moment in a point-by-point manner and averaged over the stance phase (0–100%) as

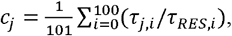

where *c*_*j*_ is the relative contribution, *τ*_*j*_ is the each moment component (j=1:resultant, j=2:GRF, j=3:rotational inertia, j=4:gyroscopic, j=5:linear inertia, j=6:gravity, and j=7:friction moment), and *τ*_*RES*_ is the resultant moment. Two-way analysis of variance (ANOVA) (moment components × foot strike conditions) with a post-hoc Tukey’s honestly significant difference (HSD) test was conducted to determine the component-wise difference of contribution *c*_*j*_ (P<0.01).

Time-series changes in joint angles, angular excursions, angular velocities, and moment component were compared between foot strike conditions using the statistical parametric mapping (SPM) two-tailed paired t-test (Pataky et al., 2013). The alpha level for the SPM test was set at 0.01. To quantify the ES of significant difference, the point-by-point Cohen d value was calculated and averaged over the phase when the SPM test detected a significant difference. Significant differences with a Cohen d value >0.8 was regarded as a large ES (Cohen, 1988). All SPM tests were implemented using the spm1d code (www.spm1d.org) in Python 3.6.3 (https://www.python.org/downloads/release/python-363/).

## Results

### Demographic and anthropometric characteristics

Participants’ mean height was 160.5 (standard deviation [SD] 5.1) cm, mean weight was 55.6 (SD 5.6) kg, and mean age was 21.2 (SD 0.9) years. Twenty-three participants used their right leg, and 2 selected their left leg.

### Approach speed

The average approach speed was not significantly different between the foot contact conditions (RFS: 1.47 [SD 0.29] m/s, FFS: 1.50 [SD 0.34] m/s, P=0.21, Cohen d=0.09). All the trials satisfied with the safety criteria of approach speed < 2.0 m/s.

### Joint moment

Two-way ANOVA demonstrated a significant main effect of moment components (hip: P<0.01, F=650.0; knee: P<0.01, F=1936.9) on the contribution to the magnitude of the resultant hip and knee moments but not the foot strike conditions (hip: P =0.75, F=0.095; knee: P=0.07, F=3.22). A significant interaction between the foot strike conditions and moment components was found in the knee (P<0.01, F=3.59) but not in the hip (P=0.15, F=1.59). For the hip and knee, the moments of GRF showed large but opposite contributions relative to the resultant moments, indicating that those 2 moment components were almost counterbalanced (Figure 3). Additionally, since the contributions of the other moment components were significantly small relative to the moment of GRFs, this study focused on the resultant moments and moments of GRF for the report of the SPM test.

**Figure 3.**
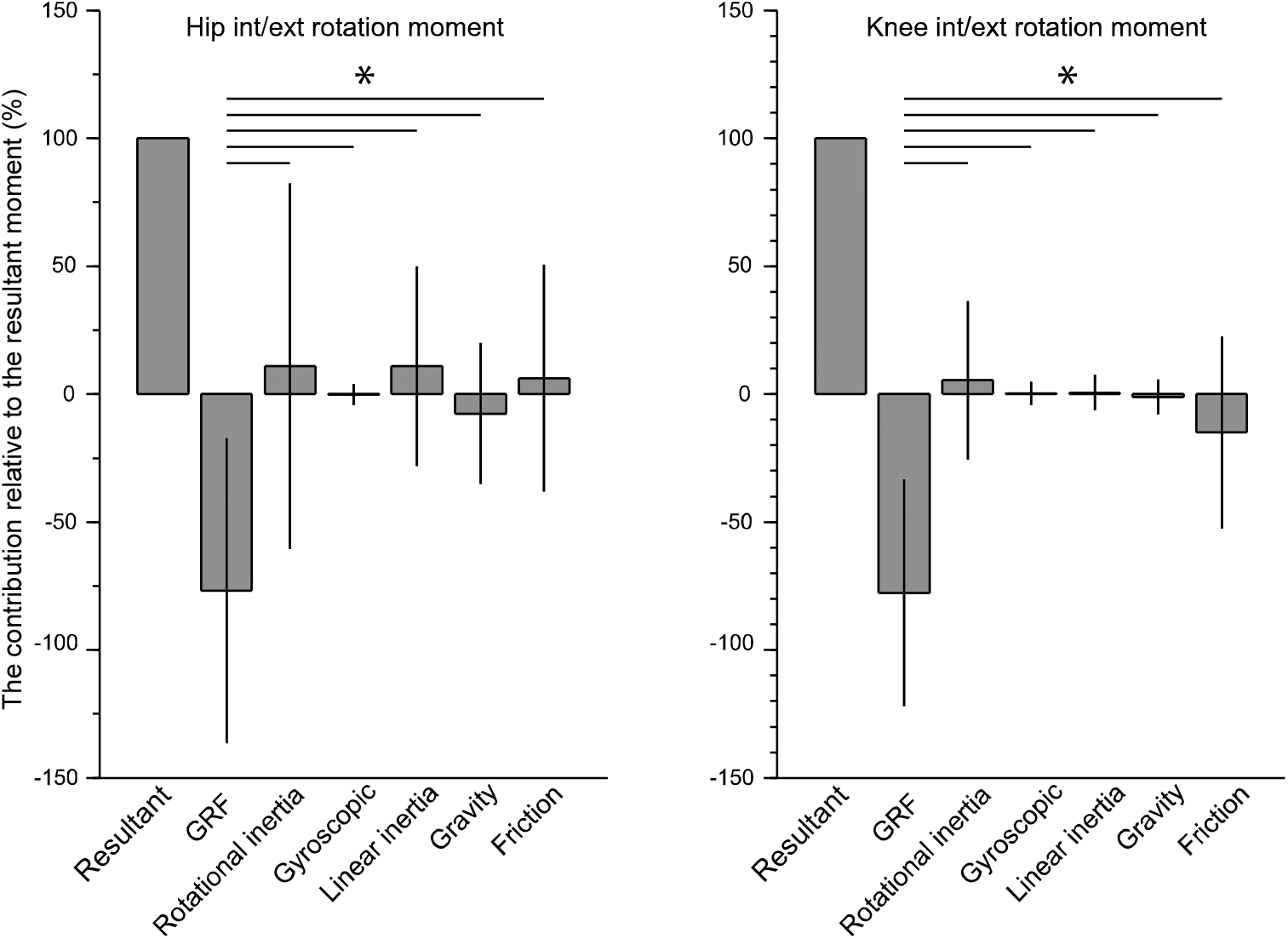
The contribution of the moment variables in the Newton-Euler equation of motion relative to the magnitude of the resultant moment. Since the two-way analysis of variance suggested that the foot strike condition has no significant effect, a post-hoc Tukey’s honestly significant difference test is used without consideration of the foot strike condition and displayed here. The results indicate that the moment of GRF has a dominant but negative contribution to the resultant moment, whereas the other moment variables show a small contribution to rotational kinetics of the hip and knee. Asterisk denotes a significant difference. GRF: ground reaction force.

For the hip and knee, the moments of GRF and resultant moments showed similar temporal patterns but in the opposite direction (Figure 4A, 4B, 4F, 4G). For the hip joint, the RFS produced a rapid increase of internally directed moment of GRF during the first 5% of the stance phase and suddenly switched to externally directed moment during the next 10–20% of the stance phase (Figure 4A). The internally directed moment of GRF at hip was significantly greater with RFS than with FFS, and it showed a very large ES (RFS: 0.12 [SD 0.03] vs. FFS: 0.01 [SD 0.00] Nm/kg, Cohen d=1.55, Figure 4A). The externally directed hip resultant moment in RFS counteracted the initial internally directed moment of GRF during the first 5% of the stance phase; however, there was no significant difference between foot strike conditions during this phase (Figure 4B). At approximately 10% of the stance phase, RFS showed significantly greater internally directed hip resultant moment than FFS did (RFS: 0.07 [SD 0.00] Nm/kg vs. FFS: −0.03 [SD 0.00] Nm/kg, Cohen d=1.29, Figure 4B).

**Figure 4.**
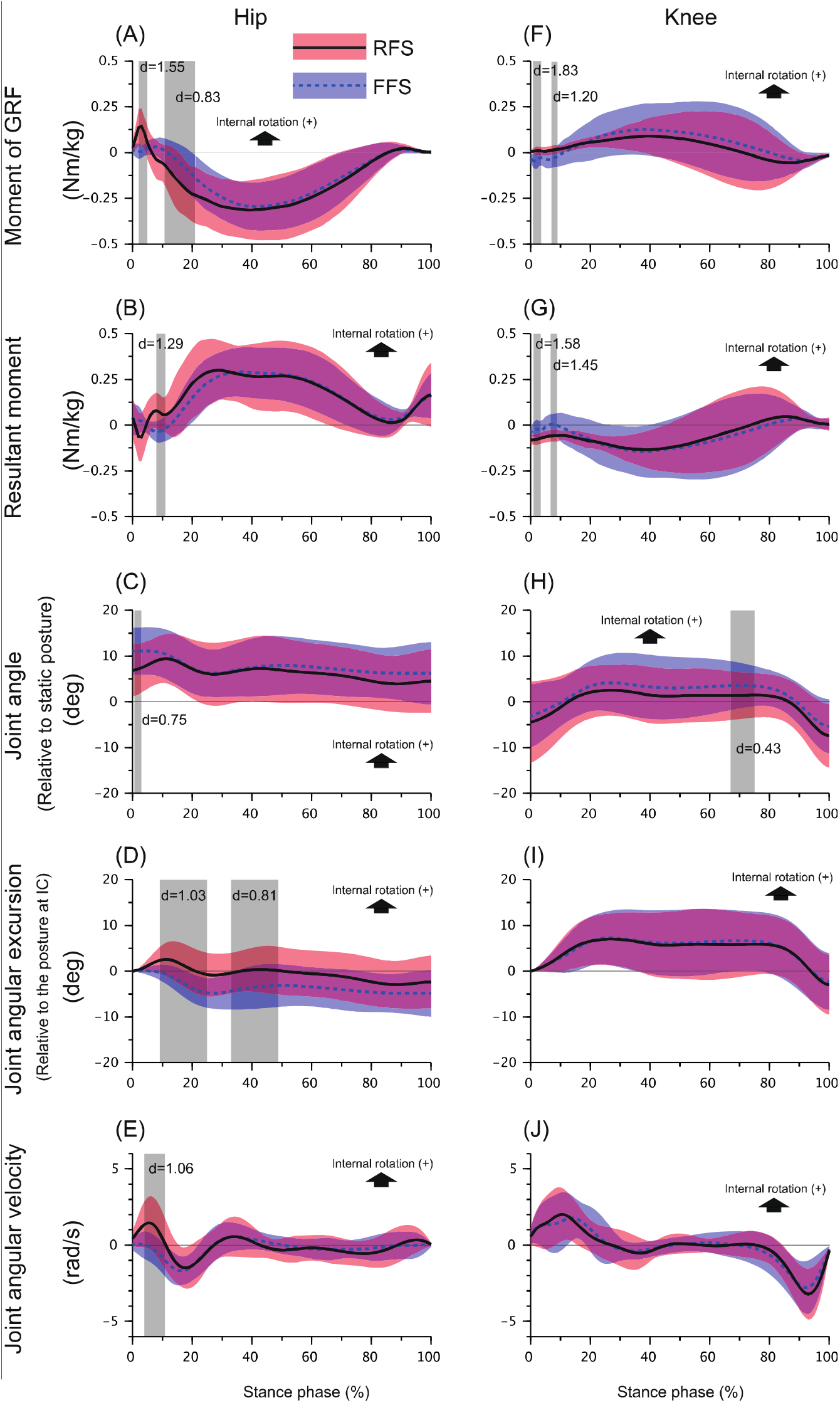
Comparison of moments, joint angles, joint angular excursions and angular velocities between foot strike patterns with SPM. Ensemble averages (standard deviations) of the moment of GRF (A, F), resultant moment (B, G), joint angle (C, H), angular displacement (D, I), and angular velocity (E, J) of the hip (left column) and knee (right column). On each panel, the solid line with orange shadow indicates the rearfoot strike condition, whereas the dashed line with the blue shadow illustrates the forefoot strike condition. The grey shaded durations are the phases where the statistical parametric mapping two-tailed paired t-test detected a significant difference with an alpha level less than 0.01. For each significant difference, the Cohen d value is calculated as an effect size. The critical thresholds are shown on each panel. (For interpretation of the references to color in this figure legend, the reader is referred to the web version of this article). GRF: ground reaction force.

Moment of GRF at the knee joint was in the opposite direction with RFS and FFS during the first 10% of the stance phase (e.g., RFS was internal, whereas FFS was external), and the ES was very large (RFS: 0.01 [SD 0.00] Nm/kg vs. FFS: −0.04 [SD 0.00] Nm/kg, Cohen d=1.83 and RFS: 0.02 [SD 0.00] Nm/kg vs FFS: −0.03 [SD 0.01] Nm/kg, Cohen d=1.20, Figure 4G). The direction of the resultant moment in both foot strike conditions were externally directed, and the RFS showed significantly greater magnitude at about 1–10% of the stance phase with large ESs (RFS: −0.07 [SD 0.00] Nm/kg vs. FFS: −0.02 [SD 0.00] Nm/kg, Cohen d=1.58 and RFS: −0.06 [SD 0.00] Nm/kg vs. FFS:0.002 [SD 0.00] Nm/kg, Cohen d=1.45, Figure 4G].

### Joint angles and Joint angle excursions

The hip was in an internally rotated position throughout the stance phase for both foot strike conditions, and there was no large ES significant difference in the hip joint angle between foot strike conditions (Figure 4C). However, the RFS caused a greater hip internal rotation excursion after IC, showing a significant difference with a large ES (RFS: 1.5 [SD1.0] deg vs. FFS: −2.14 [SD 1.20] deg, Cohen d=1.03 and RFS: 0.07 [SD 0.33] deg vs. FFS: −3.71 [SD 0.49] deg, Cohen d=0.81) during the 5–49% of the stance phase as compared to the FFS (Figure 4D). The knee showed similar rotational patterns in both foot strike conditions. There was no large ES significant difference in the knee rotational angle and excursion between the foot strike conditions (Figure 4H, 4I).

### Joint angular velocity

The RFS showed an abrupt increase of internally directed hip angular velocity after IC, which was significantly higher than that of FFS during the 4–11% of the stance phase (RFS: 1.21 [SD 0.23] rad/s vs. FFS: −0.18 [SD 0.24] rad/s, Cohen d=1.06, Figure 4E). The knee angular velocities in both foot strike conditions were commonly internally directed during the first 30% of the stance phase and externally directed during the last 20% of the stance phase, with no significant difference between the conditions (Figure 4J).

## Discussion

This study is the first to report that the foot strike pattern differentiated the horizontal-plane hip and knee kinetics and kinematics during the early stance phase of a cutting maneuver in female athletes. Specifically, the RFS produced a rapid increase of internally directed moment of GRF at the hip during the first 5% of the stance phase, subsequent increase of the hip internally directed joint angular velocity during 4–11% of the stance phase, and hip internal rotational excursion peaking at approximately 10% of the stance phase (Figure 4A,E,D). Those characteristics were not found in the FFS condition. The critical difference found in the knee kinetics was that the RFS and FFS demonstrated the opposing directions of the moment of GRF during 1–10% of the stance phase, i.e., the RFS produced the internally directed while the FFS showed the externally directed moment of GRF (Figure 4F, G). For both hip and knee joint, the moments of GRF were the significant determinants of the resultant joint moments (Figure 3), providing insight into the generation of the high-risk hip and knee loading patterns during the cuttings in females. Generally, those findings supported our hypotheses.

The key finding of this study was that the hip and knee loading patterns (direction of the moment of GRF at hip and knee) was different between foot strike pattern and this difference may explain the discrepancy in the ACL injury risk associated with the foot strike pattern. The RFS exhibited the combination of internal directed hip and knee moments of GRF at first 10% of stance (Figure 4A, F), and the hip and knee internal rotation excursions (Figure 4D,I). Contrary to this, the FFS showed the combination of externally directed hip and knee moments of GRF at the same time frame (Figure 4A, F). The increased hip internal rotation excursion in female athletes during the functional movements has been interpreted as a risk factor for ACL injury (McLean et al., 2005; Pollard et al., 2007; Sigward et al., 2015) because the hip internal rotation in weight bearing situation results in the subsequent the knee valgus kinetic chain (McLean et al., 2005). In addition, the internal tibial rotation moment has been reported to increase the in-situ force on the ACL (Kiapour et al., 2016) and is associated with the mechanism of ACL injury (Koga et al., 2010; Shimokochi and Shultz, 2008), whereas the tibial external rotation has opposite mechanical effect on ACL (Markolf et al., 1995). In this study, even in a slow approach speed (< 2.0 m/s) relative to the previous cutting studies (Sigward et al., 2015; Vanrenterghem et al., 2012), the significant differences of hip and knee rotational GRF loading pattern with the opposite directions were found between the foot strike patterns. Those directional discrepancy in the GRF loading pattern is expected to expand in a more demanding cutting. Therefore, from the perspective of the horizontal-plane hip and knee kinetic chain, it is suggested that the RFS potentially add the risk of ACL injury whereas the FFS is relatively an ACL-protective foot landing technique.

The initial increase of hip internal rotation angular velocity in the RFS was obviously caused by the prominent increase of the internally directed moment of GRF at the hip (Figure 4A,E). In a more demanding situation, the effect of this GRF loading would change the orientation of the entire stance limb relative to the direction of travel at early phase. In such a stance limb configuration, the knee extension/flexion axis will no longer maintain an appropriate orientation along the GRF line of action when the full weight acceptance phase, resulting in a risky knee loading patterns such as the combined knee valgus and tibial internal rotation loading (Ogasawara et al., 2020). In this sense, the initial hip internal rotation in the early phase of RFS may be a precursor mechanism developing the subsequent knee misorientation relative to the GRF vector and may change the knee kinetics.

In this study, the hip internal rotation excursion in RFS showed a temporal increase peaking around 10 % of stance but it did not maintain over the stance phase (Figure 4D). Similarly, the temporal hip internal rotation at early stance phase was consistent among the lab-controlled cutting studies which the ACL injury was not observed during their experiments (Havens and Sigward, 2015b; Imwalle et al., 2009). This implied that, in the previous lab-controlled studies as well as this study, the magnitude of hip internal rotation moment due to GRF was not large enough to develop a complete hip internal displacement which was sufficient to threaten the ACL. The quantitative video analyses of the actual ACL injury event have demonstrated that the initial hip internal rotation continued until the ACL injury occurred (Koga et al., 2018). In contrast, the video analysis of the high-risk motion with non-injury situation demonstrated no prominent hip internal rotations within 40 ms after IC as well as the knee valgus and tibial internal rotation (Sasaki et al., 2017). Therefore, it may be speculated that the presence of the complete hip internal rotation at the beginning of the stance phase determines the subsequent stance limb configuration and affects the risk of ACL injury.

Limited hip internal rotation range of motion (RoM) has been reported to be associated with the incidence of noncontact ACL injury (Boutris et al., 2018) and rerupture of the ACL (Gomes et al., 2014). This information seems to be contradictory to our aforementioned proposal. However, we believe that those clinical findings and our proposal do not conflict because the time scopes were different. As aforementioned, we suggested that the initial hip internal rotation due to internally directed GRF moment acting at hip may be problematic for subsequent alignment control; therefore, this concern would be pronounced in the pre-injury phase (e.g., period from IC to ligamentous rupture). Contrary to this, a narrow hip rotational RoM may become problematic at about the terminal of hip RoM since cadaveric studies demonstrated that the knee rotational stress increases as the hip rotation is mechanically constrained by the limit of RoM (Beaulieu et al., 2014). Hence, these mechanics may occur around the “rupture phase” of ACL injury. It could be also speculated that if the narrow hip internal rotation RoM reflects the dysfunction of hip external rotator muscles, such as contracture by muscle fatigue, it may not adequately counteract the internally directed GRF loading acting at hip at the foot impact phase. However, the effect of passive structures, such as an increased femoral alpha angle in the anteroposterior view (VandenBerg et al., 2016), could also contribute to the limited hip internal RoM and be associated with noncontact ACL injury; therefore, the dysfunction of hip external rotators alone does not fully explain the association between the static measure of hip rotation RoM and the mechanism of ACL injury. Further study about the relationship between the static hip RoM and dynamic hip and knee kinetic chain during functional activities will be needed to clarify this issue.

The component-wise analysis of the equation of motion demonstrated that the externally directed hip resultant moment (muscle moment) counteracted the internally directed moment of GRF exerted at the hip (Figure 4A, B), suggesting that adequate hip muscular control was essential to maintain a neutral hip orientation after IC of the cutting maneuver. This was also supported by our results that the magnitudes of the other moment components were too small to balance the moment of GRF, suggesting that the hip muscle moment was a primary counterpart of the moment of GRF (Figure 3). If the hip rotator muscles failed to increase the horizontal-plane joint impedance prior to foot impact and could not adequately balance the internally directed moment of GRF exerted at the hip, the initial hip internal excursion would become more pronounced. Leetun et al.(LEETUN et al., 2004) reported that female athletes had significantly decreased hip external rotation isometric strength (percentage body weight) than male athletes, and they prospectively revealed that hip external rotation strength was the effective predictor of the likelihood of sustaining noncontact injuries, including ACL rupture. Omi et al. (Omi et al., 2018) suggested the use of a “hip-focused” training to increase hip rotational strength and hip neuromuscular control; they achieved 62% reduction of noncontact ACL injuries in female basketball players. Collectively, these clinical findings highlight the importance of hip external rotator strength for managing the risk of ACL injury (McLean et al., 2005). Our results support the mechanical rationale underlying those previous findings regarding how hip external rotator moments contribute to the control of hip rotational kinematics during athletic maneuvers.

The limitation of this study was that the current results were from female participants, so caution is needed when generalizing our findings to male athletes. Sex disparities are known to exist in the hip and knee biomechanics during cutting maneuvers (Pollard et al., 2007); thus, this study used female participants to isolate the effect of foot strike pattern. The slow approach speed could be a limitation of this study from the perspective of the small magnitudes of the vector quantities. However, the temporal patterns of the hip and knee kinetics and kinematics were consistent with results of the previous similar cutting studies (Pollard et al., 2004), suggesting the conclusion of this study regarding the GRF loading patterns (direction of the GRF moments on the hip and knee) was not affected by the slow approach speed.

## Conclusion

In conclusion, the RFS showed a greater hip internal rotation angular velocity combined with knee internal rotation during the early phase of the cutting task which was not seen in FFS. Such a hip and knee kinetic combination in the RFS would potentially increase the risk of ACL injury in female athletes. Hence, FFS is recommended as a safer cutting strategy than RFS in female athletes.

## Supporting information

Supplemental Content

## Data Availability

Data that support the findings of this study will be made available from the corresponding author upon reasonable request.

## Acknowledgements

The authors acknowledge support from Chisa Aoyama, Mai Kitaura and Yui Kawano for the assistance in data collection. All authors have no conflicts of interest related to this study. The authors declare that the results of this study are presented clearly, honestly, and without fabrication, falsification, or inappropriate data manipulation.

## Funding

This study was supported by the Grant-in-Aid for Young Scientists (B) project [grant number: 24700716] funded by the Ministry of Education, Culture, Sports, Science and Technology (MEXT), Japan.

## Contributions

I.O., Y.S., S.K., and K.N. conceptualized and designed the study.

I.O. organized the experiments.

I.O. performed the experiments and data collection.

I.O., Y.S., and S.K. analyzed the data and performed the statistical analysis. I.O., Y.S., S.K., and K.N. wrote the manuscript.

I.O., Y.S., S.K., T.M., and K.N. reviewed the manuscript, suggested corrections and approved its final version.

## Notes

**Conflict of interests:** None.

### Competing Interest Statement

The authors have declared no competing interest.

### Author Declarations

Mukogawa Women's University, No. 12-13

