## Supplemental Content for "Effect of rearfoot strikes on the hip and knee rotational kinetic chain during the early phase of cuttings in female athletes"

**Supplement Content**

***Definition of the kinematic model***

The kinematic model consisted of four segments (
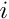
=1: foot,
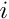
=2: shank,
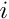
=3: thigh, and
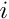
=4: pelvis) with three joints (
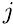
=1: ankle,
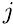
=2: knee, and
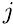
=3: hip) (Figure s1). Each joint center was modeled as a ball joint of 3 degrees of freedom and expressed as a point
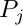
 (
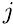
=1, 2, 3) in the global coordinate system (GCS). The point
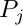
 corresponded with the proximal end of the segment
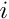
 and the distal end of the segment
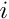
+1. The position vector from the origin of the GCS to the point of each joint center
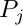
 was defined as
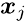
 (
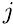
=1, 2, 3), and the position vector going to the segmental center of gravity (CoG) was defined as
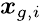
 (
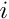
=1, 2, 3). The local coordinate system (LCS) of segment
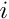
 was defined as a 3×3 rotation matrix,
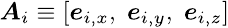
, consisting of three common perpendicular unit base vectors:
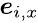
 was the anterior/posterior axis pointing forward,
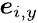
 was the vertical axis along with the longitudinal axis of the segment pointing upward, and
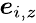
 was the medial/lateral axis pointing to the right of segment
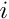
. The origin of the LCS was fixed at the CoG of each segment.
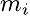
,
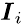
, and
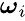
 were the mass, inertia tensor, and segmental angular velocity vector of segment
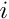
, respectively. The ground reaction force (GRF) vector
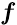
 and friction torque vector
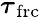
 were acting at the CoP of the foot segment. Note that the friction torque vector
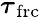
 had the only vertical component around the CoP. Vector
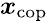
 was the position vector going from the origin of the GCS to the CoP. Vector
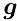
 denotes the gravitational acceleration.

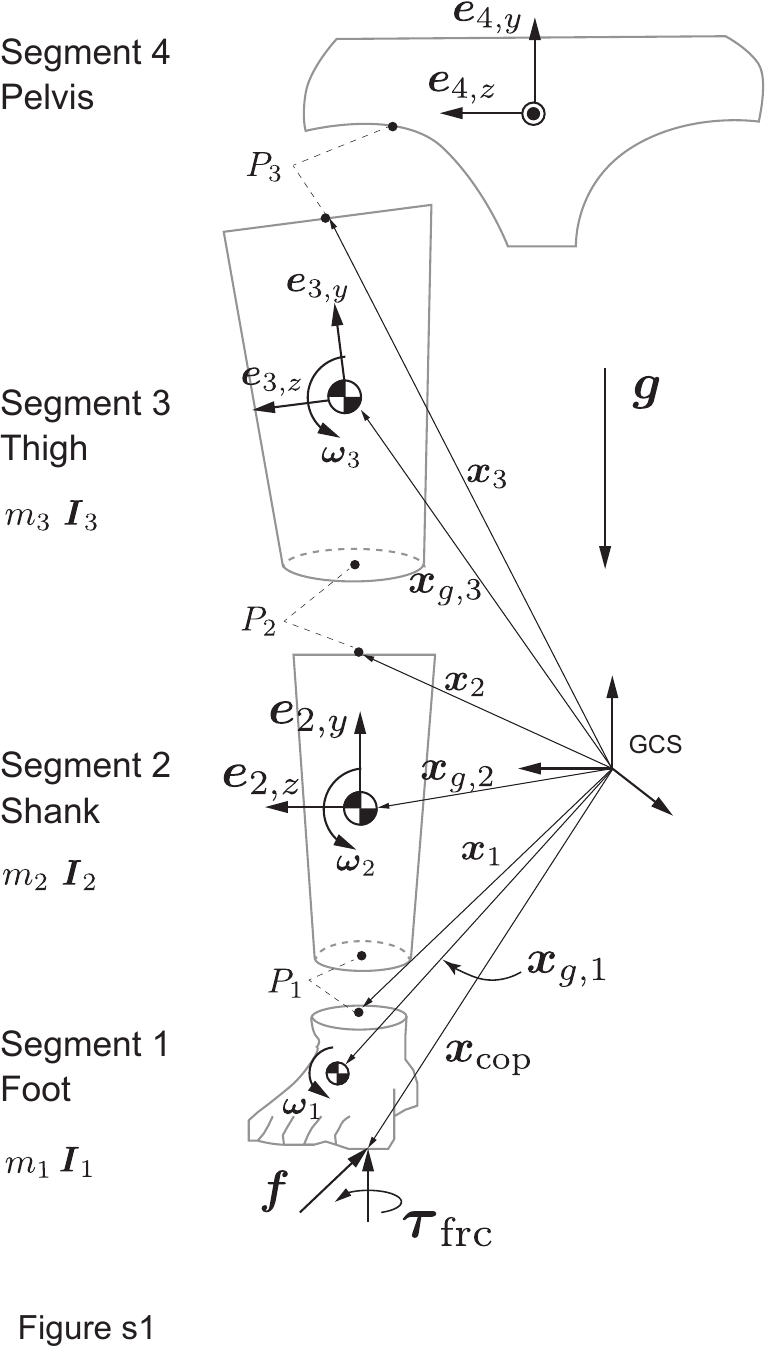

*Figure 1s. Three-dimensional four-link kinematic model*

***Joint angle calculation***

For the rotational joint angle calculation, we referred to the concept of the joint coordinate system (1, 2). This concept is used to calculate a unit floating vector that is mutually perpendicular to the vertical axis (y) of the distal segment and medial/lateral axis (z) of the proximal segment:

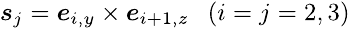
. (1)

This unit floating vector is always facing anterior of the proximal segment while keeping orthogonal to the longitudinal axis of the distal segment. Using this unit floating vector, the hip and knee internal/external rotation angle was calculated as follows:

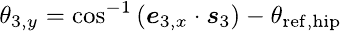
 (2)

and

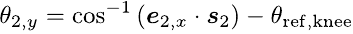
 (3)

, where
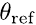
 is the reference angle to offset.

***Newton-Euler equation of motion for inverse dynamics***

The intersegmental torque vector
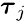
 acting about the hip (
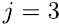
) and knee (
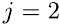
) joint was calculated as the resultant sum of the rotational inertia torque, gyroscopic torque, moment of linear inertia force, moment due to gravity, moment of GRF, and moment due to friction torque:

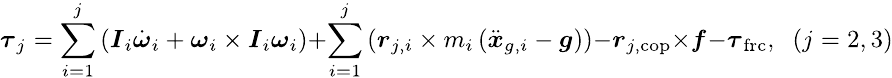
 (4)

, where

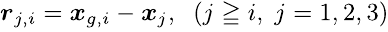
 (5)

is the moment arm vector going from joint j to the CoG of the distal segment and

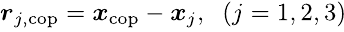
 (6)

is the moment arm vector going from joint j to the CoP of the foot segment. In the typical recursive procedure, the equation of motion was usually formulated around the CoG of the segment and calculated in a segment-by-segment manner (3). However, Eq.(4) was developed around the joint center of interest in order to consider the effect of all external forces and moments that potentially alter the mechanical state of the system (Figure s2).

The internal(+)/external(-) rotation torque component of each torque variable in Eq.(4) was obtained by taking a dot product of the given torque variable vector with the unit base vector of the vertical axis of the segment. For example, the hip internal/external rotation components of the intersegmental torque and moment of GRF were as follows:

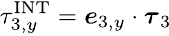
 (7)

and

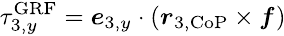
. (8)

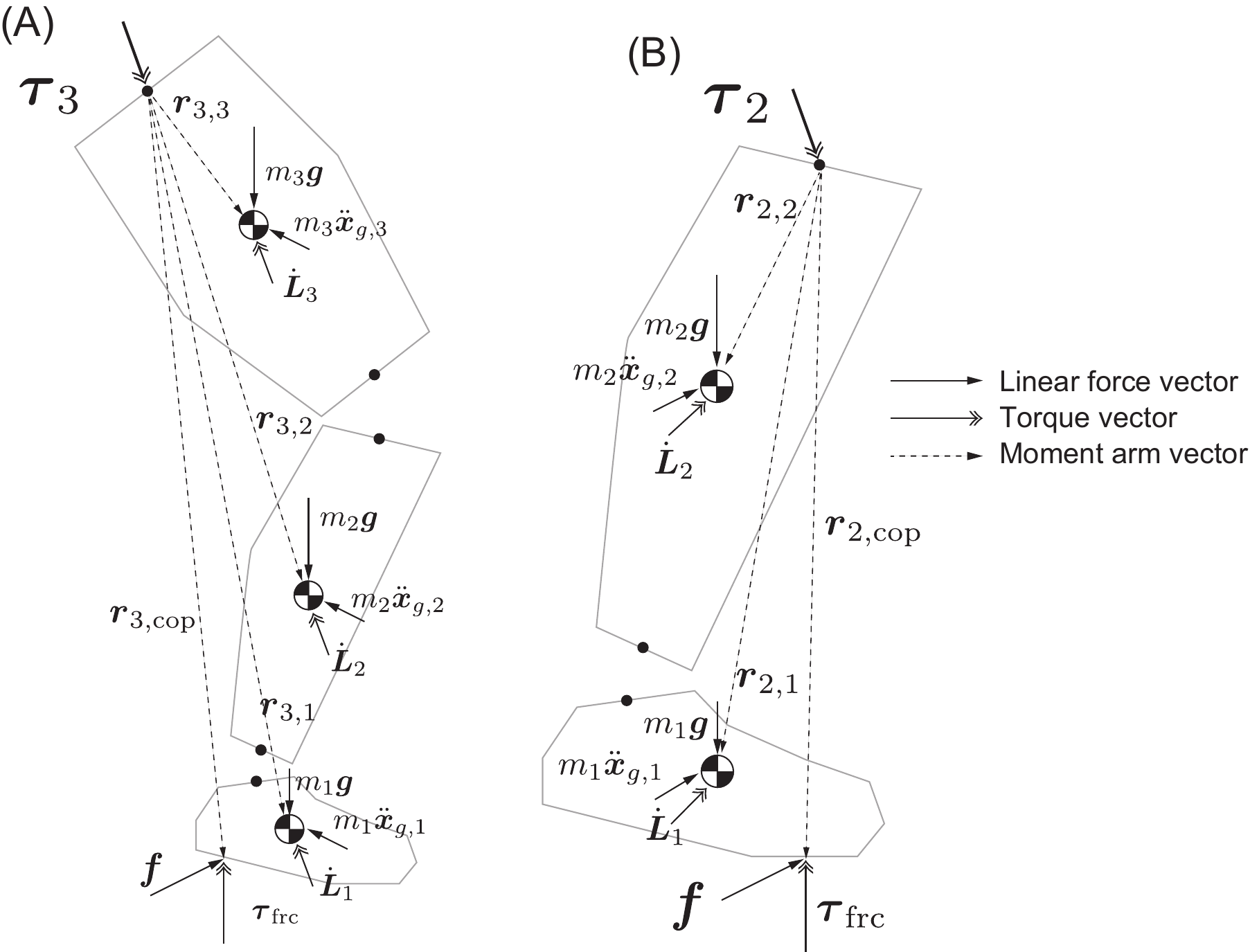

*Figure s2. Free body diagrams for the calculations of (A) hip and (B) knee intersegmental torques.*

Note that
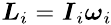
. The dashed arrows represent the moment arm vector from the joint center of interest to the acting points of the linear forces, such as gravity, linear inertia acting at the CoG of the segment, and the GRF acting at the CoP of the foot segment.
